# Impact of a nutrition-sensitive agroecology program in Andhra Pradesh, India, on dietary diversity, nutritional status, and child development

**DOI:** 10.1101/2023.05.16.23290036

**Authors:** Lakshmi Durga, Yandrapu Bharath, Lilia Bliznashka, Vijay Kumar, Veerendra Jonnala, Vijayalakshmi Chekka, Srileka Yebushi, Aditi Roy, Nikhil Srinivasapura Venkateshmurthy, Poornima Prabhakaran, Lindsay M. Jaacks

**Affiliations:** Rythu Sadhikara Samstha, Guntur, Andhra Pradesh, India; Public Health Foundation of India, New Delhi, India; Global Academy of Agriculture and Food Systems, The University of Edinburgh, Midlothian, UK; International Food Policy Research Institute, Washington DC, USA; Centre for Chronic Disease Control, New Delhi, India; Harvard TH Chan School of Public Health, Boston, Massachusetts, USA

**Author notes:** Corresponding author: (LB).

**Keywords:** Nutrition-sensitive agriculture, maternal and child health, sustainable consumption, agroecology, home gardens, diet quality, child development

## Abstract

**Introduction:** To date, most food-based nutrition interventions have not considered food production practices, particularly the use of synthetic chemicals. This study aimed to evaluate the impact of a multi-component food-based nutrition intervention involving homestead food production, nutrition counselling, cooking demonstrations, and crop planning exercises, and employing agroecological production practices (herein ‘nutrition-sensitive agroecology program’), on dietary diversity, nutritional status, and child development in Andhra Pradesh, India.

**Methods:** A cross-sectional assessment was conducted in 2021-2022 of 50 intervention villages where the nutrition-sensitive agroecology program had been implemented since 2018 and 79 control villages where only the agroecology program had been implemented. Data on self-reported dietary intake, caregiver-reported early child development, anthropometric measurements, and hemoglobin concentrations were collected using standardized procedures by trained Nutrition Farming Fellows, who were also responsible for implementing the program.

**Results:** A sample of 3,511 households (1,121 intervention and 2,390 control) participated in the survey. Dietary diversity scores (DDS) among women and men were mean (SD) 6.53 (±1.62) and 6.16 (±1.65), respectively, in intervention villages and 5.81 (±1.58) and 5.39 (±1.61), respectively, in control villages (p<0.01). DDS among children 6-24 months of age in intervention and control villages was 2.99 (±1.52) and 2.73 (±1.62), respectively (p<0.01). Children <2 years of age were less likely to be anemic in intervention versus control villages (59% versus 69%, p<0.01). Children 18-35 months age in intervention villages had higher child development scores than children in control villages (all p<0.05).

**Conclusion:** Nutrition-sensitive agroecological programs may be effective in improving diets, nutrition, and child development in rural India.

## Introduction

India, which is home to one-sixth of the global population, ranks 107th on the Global Hunger Index and suffers from a high burden of undernutrition (1) despite economic development (2). According to the latest National Family Health Survey (NFHS, 2019-21), 35.5% of children <5 years of age are stunted, 19.3% are wasted, and 57.0% of reproductive-aged women are anemic (3). These numbers reflect only small improvements in stunting and wasting, and a slight worsening of anemia compared to the previous round of NFHS, conducted in 2015-16, which found child stunting, wasting, and women’s anemia were 38.4%, 21.0%, and 55.3%, respectively (4).

Successive governments in India have attempted various interventions aimed at addressing undernutrition. Most recently, food-based strategies to promote dietary diversity have been highlighted by the Government of India, including in the Poshan (nutrition) 2.0 guidelines by the Ministry of Women and Child Development (5). To date, the most common food-based strategy employed has been nutri-gardens, sometimes referred to as kitchen gardens or homestead gardening. Several evaluations of nutri-gardens have been conducted in India (6–12). For example, one study in Telangana in south India evaluated a home garden and backyard poultry intervention coupled with nutrition education and found a significant increase in the quantity of green leafy vegetables and eggs consumed by women and children (10). Beyond India, evidence from other South Asian countries and Sub-Saharan Africa shows that nutri-gardens are an effective nutrition-sensitive agricultural intervention to improve women and children’s diets through the production of nutrient-dense products and dietary diversification (7,13).

Most nutrition-sensitive agricultural interventions to date have not emphasized agroecological production methods, including limiting the use of synthetic chemicals such as pesticides. However, given increasing evidence of adverse effects of pesticides on child growth (14) and development (15), the production practices used for nutri-gardens are an important consideration. Moreover, most evaluations of nutri-garden interventions have focused on the impact on women and children’s dietary diversity, or, in a few cases, on women and children’s nutritional status including anthropometry and anemia (7). Important gaps remain in understanding the impact of nutrition-sensitive agricultural interventions on under-studied outcomes including child development, men’s dietary diversity, and women’s anemia.

The objective of this study was to evaluate the impact of a government funded and implemented nutrition-sensitive agroecology program on adult and child dietary diversity, nutritional status, and child development. Our study advances the understanding of the importance of pesticide-free production practices in the context of food-based strategies to improve nutrition and child development.

## Methods

### Study context

The study was conducted in the south Indian state of Andhra Pradesh, known as the ‘rice bowl of India’ where 63% of households depend on agriculture and 37% of land area is under agriculture (including fishponds) (16). The main nutrition problems are child stunting (31%), child wasting (16%), and women’s anemia (59%) (17). Rates of non-communicable diseases in Andhra Pradesh, particularly diabetes and hypertension, are also on the rise (17).

### Intervention

This study evaluated a food-based nutrition intervention that was embedded within a larger government program, called the Andhra Pradesh Community managed Natural Farming (APCNF), previously referred to as Zero Budget Natural Farming (ZBNF). APCNF is an ongoing community-based transformation program implemented by Rythu Sadhikara Samstha (RySS), a not-for-profit company established by the Government of Andhra Pradesh. The program aims to convert all six million farmers and six million hectares of land to natural farming by the year 2031 (18).

The APCNF intervention, conceived as a climate-resilient, agroecological intervention, comprises use of locally sourced natural inputs to act as bio stimulants to the soil. APCNF is grounded in nine universal principles: (1) soil to be covered with crops 365 days (living root principle), (2) diverse crops, 15–20 crops, including trees, (3) keep the soil covered with crop residues, whenever living plants are not there, (4) minimal disturbance of soils, minimize tillage, (5) farmers’ own seeds or indigenous seeds, (6) integrate animals into farming, (7) bio stimulants as catalysts to trigger soil biology, (8) pest management through better agronomical practices and botanical pesticides, and (9) no synthetic fertilizers, pesticides, herbicides, or weedicides. The intervention is implemented through women collectives known as self-help groups and their federations (19).

The food-based nutrition intervention comprised a variety of initiatives to nudge households towards better dietary diversity, health seeking behaviors, and nutrition. Specific initiatives included implementing homestead food production (nutri-gardens, backyard poultry and small ruminants, and fisheries), nutrition counselling at Farmer Nutrition Schools, cooking demonstrations, crop planning exercises, and behavior change communication (BBC). Participatory Learning and Action tools were employed to counsel women attending the Farmer Nutrition Schools. The cooking demonstrations involved recipe preparation with locally available ingredients and cooking methods for retention of nutrients.

The target beneficiaries consisted of pregnant and lactating women, mothers of children under 2 years of age, adolescent girls, and women from the ‘poorest of the poor; households. The ‘poorest of the poor’ households included agricultural labor households that did not own land, nor take land on lease, or belonged to Scheduled Caste / Scheduled Tribe communities and owned less than two acres of land (19). The Scheduled Caste / Scheduled Tribe communities are the most marginalized sections of society.

These interventions were implemented by Nutrition Farming Fellows (NFFs) trained in Home Science, whose primary objective was to counsel the target beneficiaries to encourage nutritious food consumption behaviors. The NFFs resided in their allocated villages, for which they were paid by RySS. The NFFs worked in synergy with various community programs such as Integrated Child Development Scheme, that provides an array of services to children below 6 years and their mothers. Beginning at the end of 2018, 50 APCNF villages received this food-based nutrition intervention (herein ‘intervention villages’). Villages were selected from each of the state’s 13 districts, as shown in Fig 1 (as of April 2022, there were 26 districts in Andhra Pradesh).

**Figure 1.**
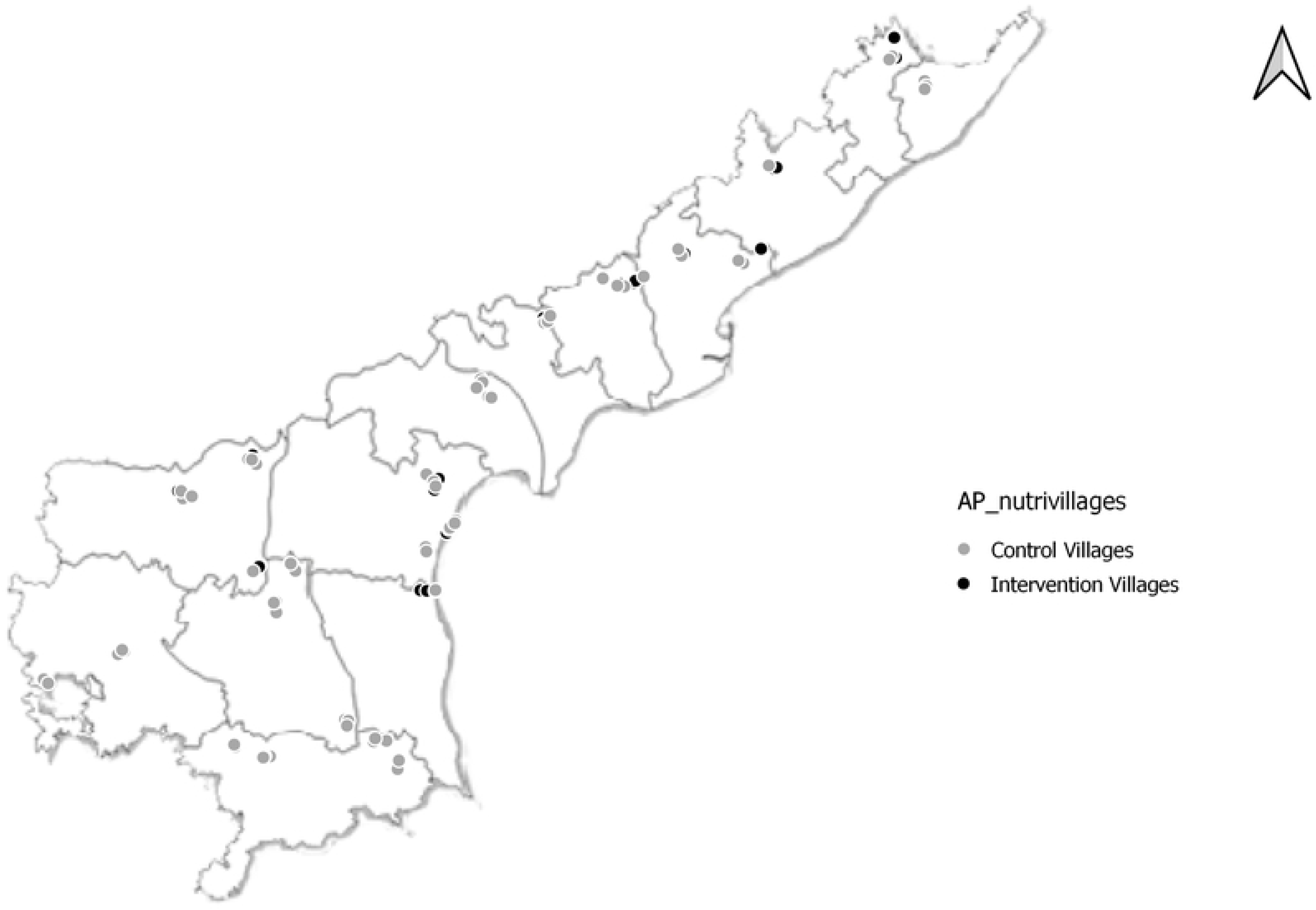
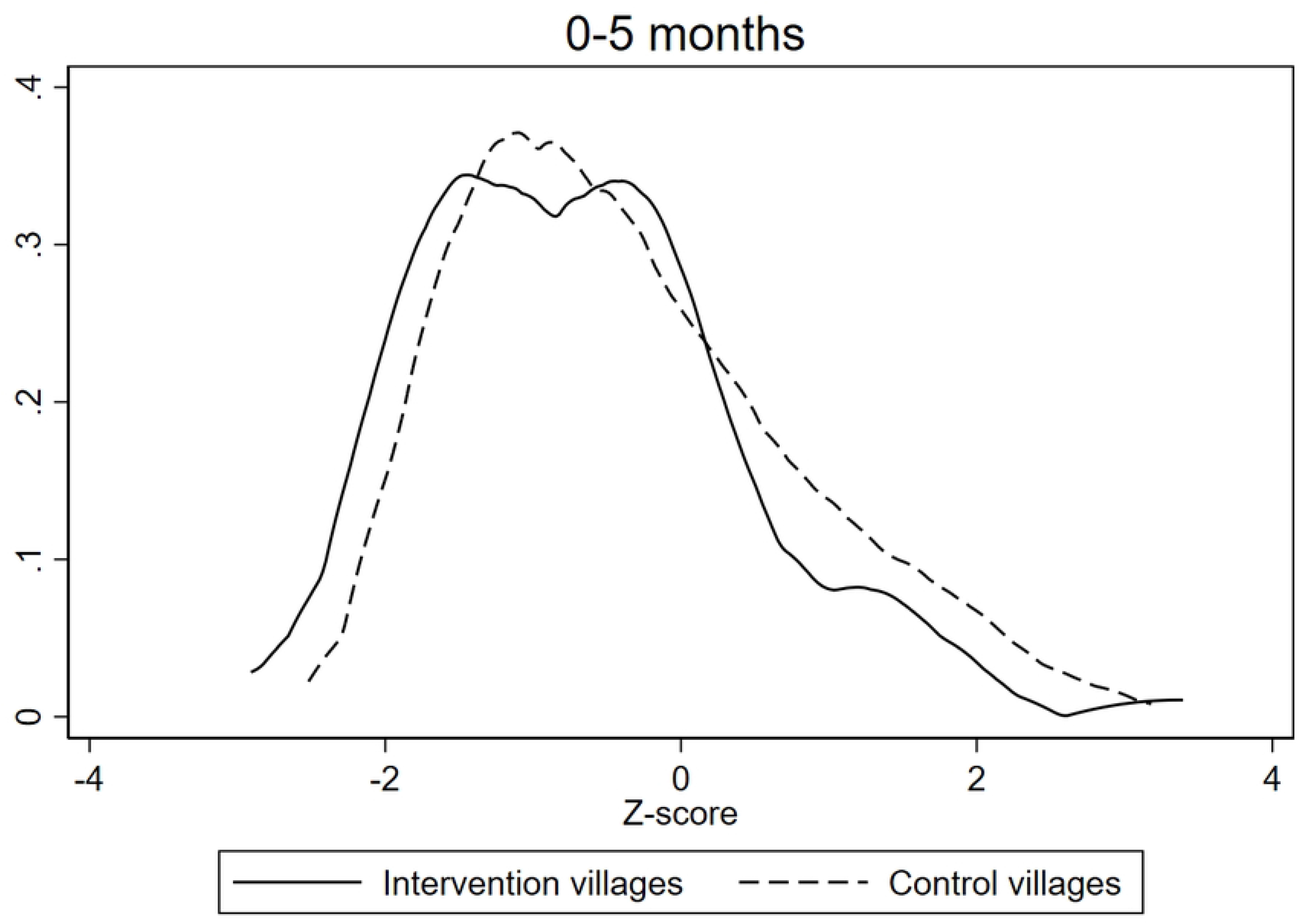
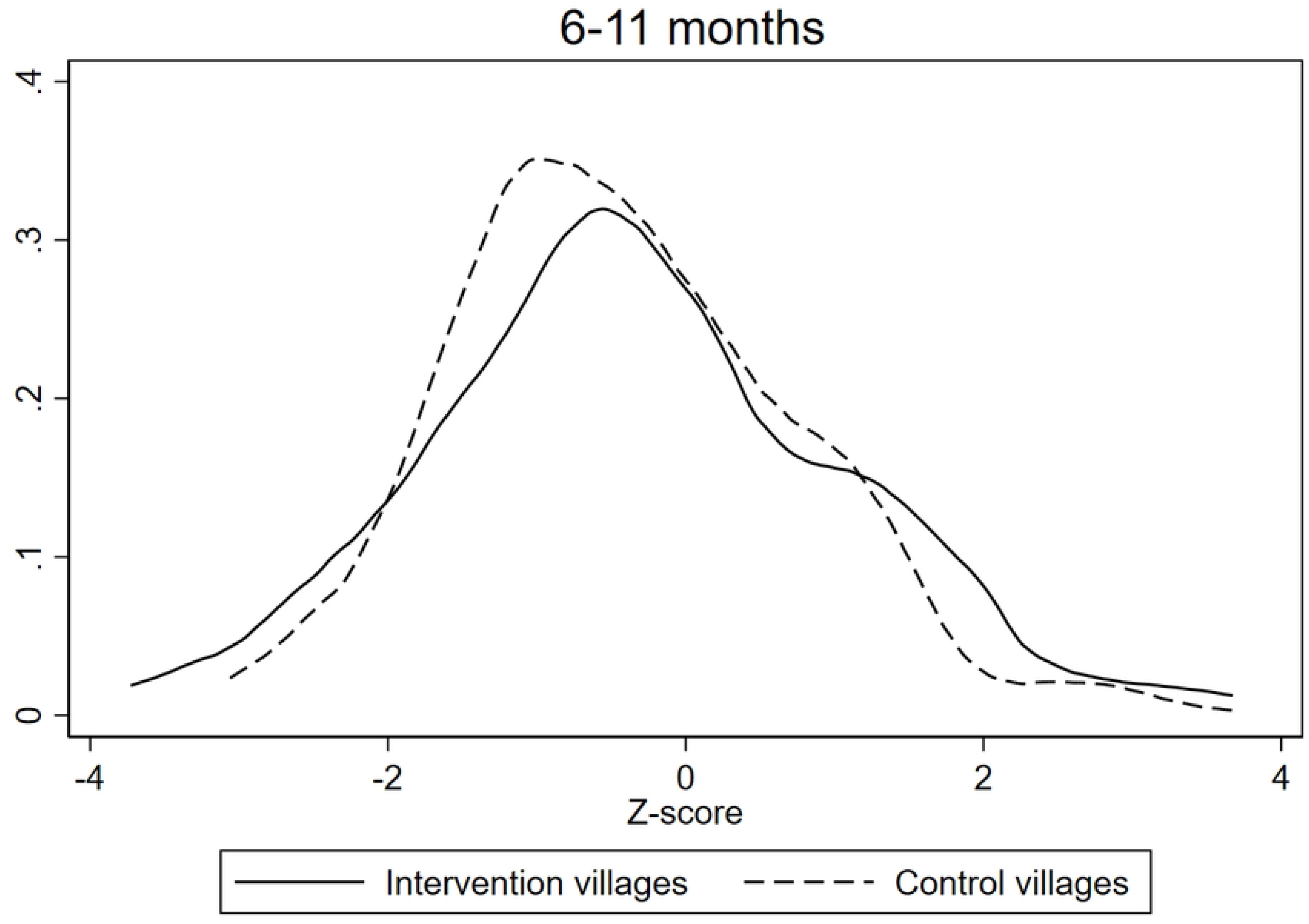
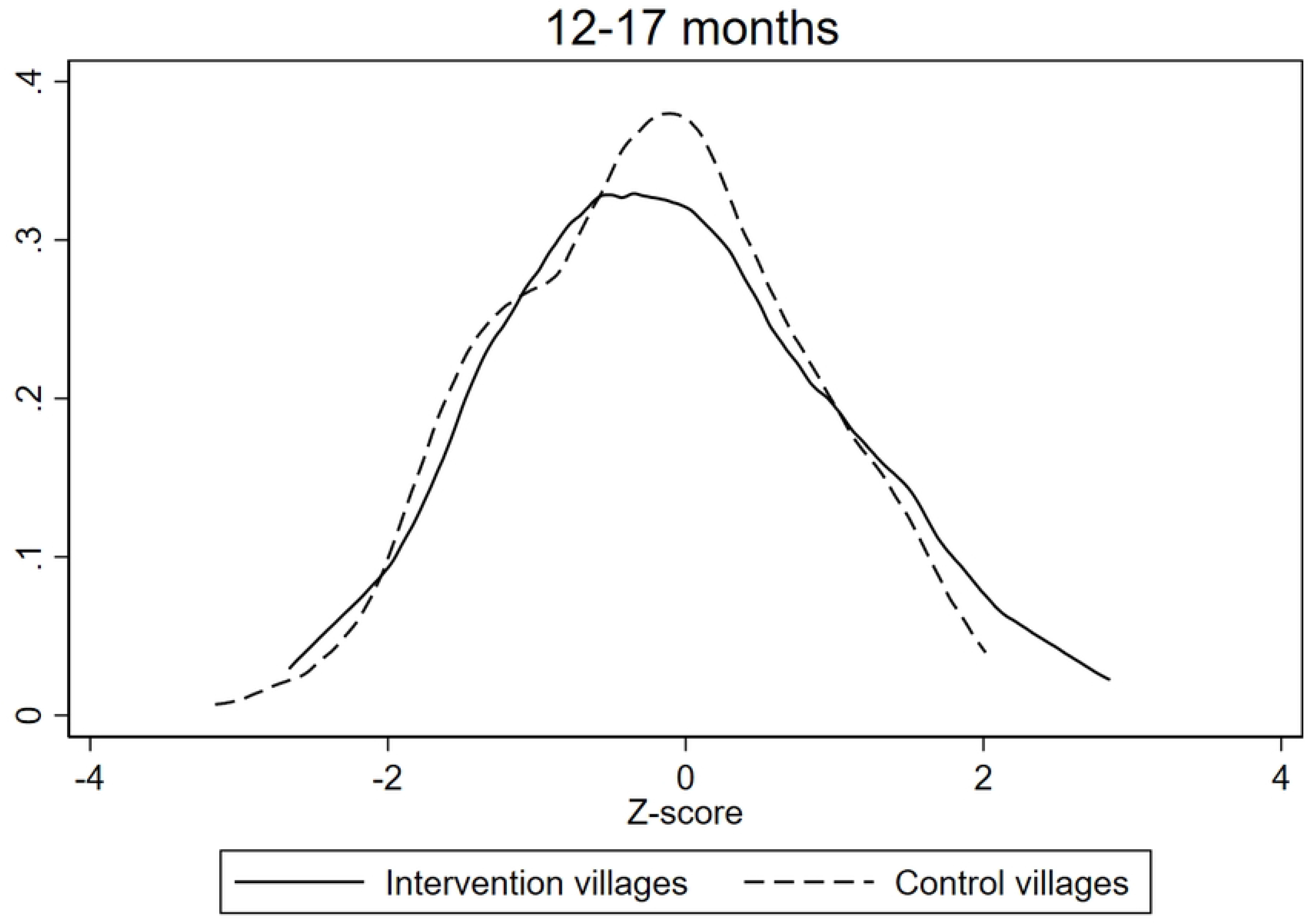
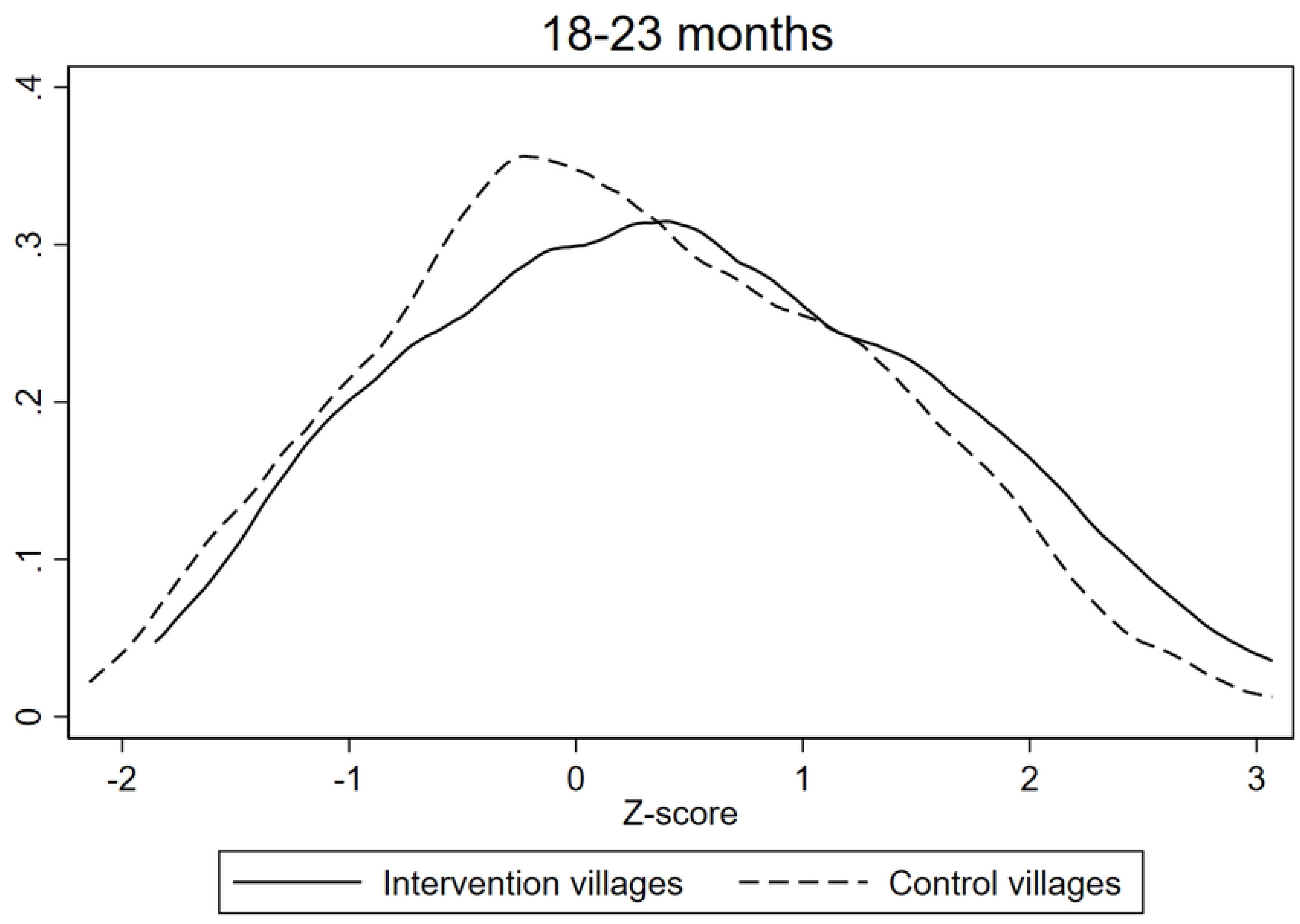
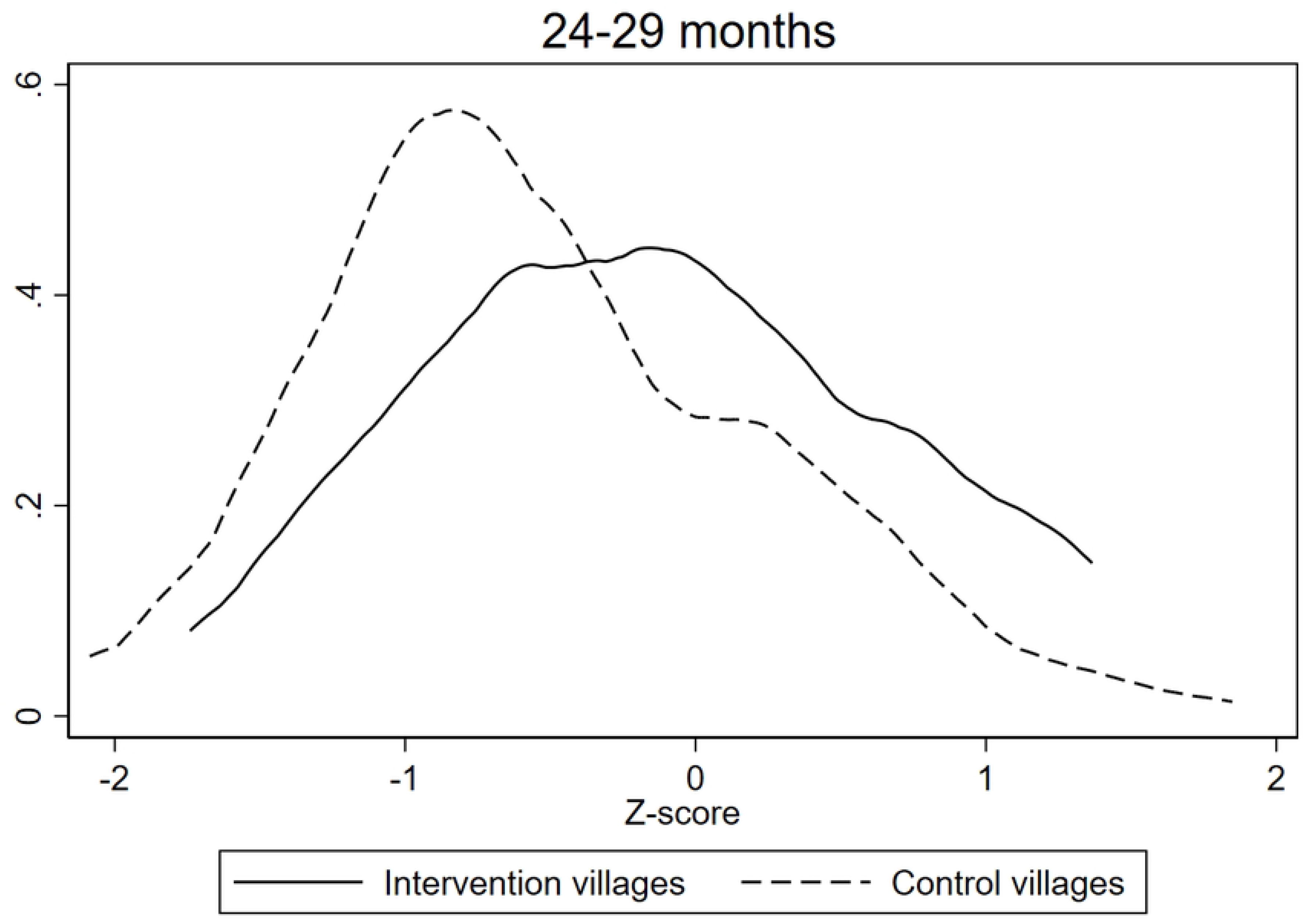
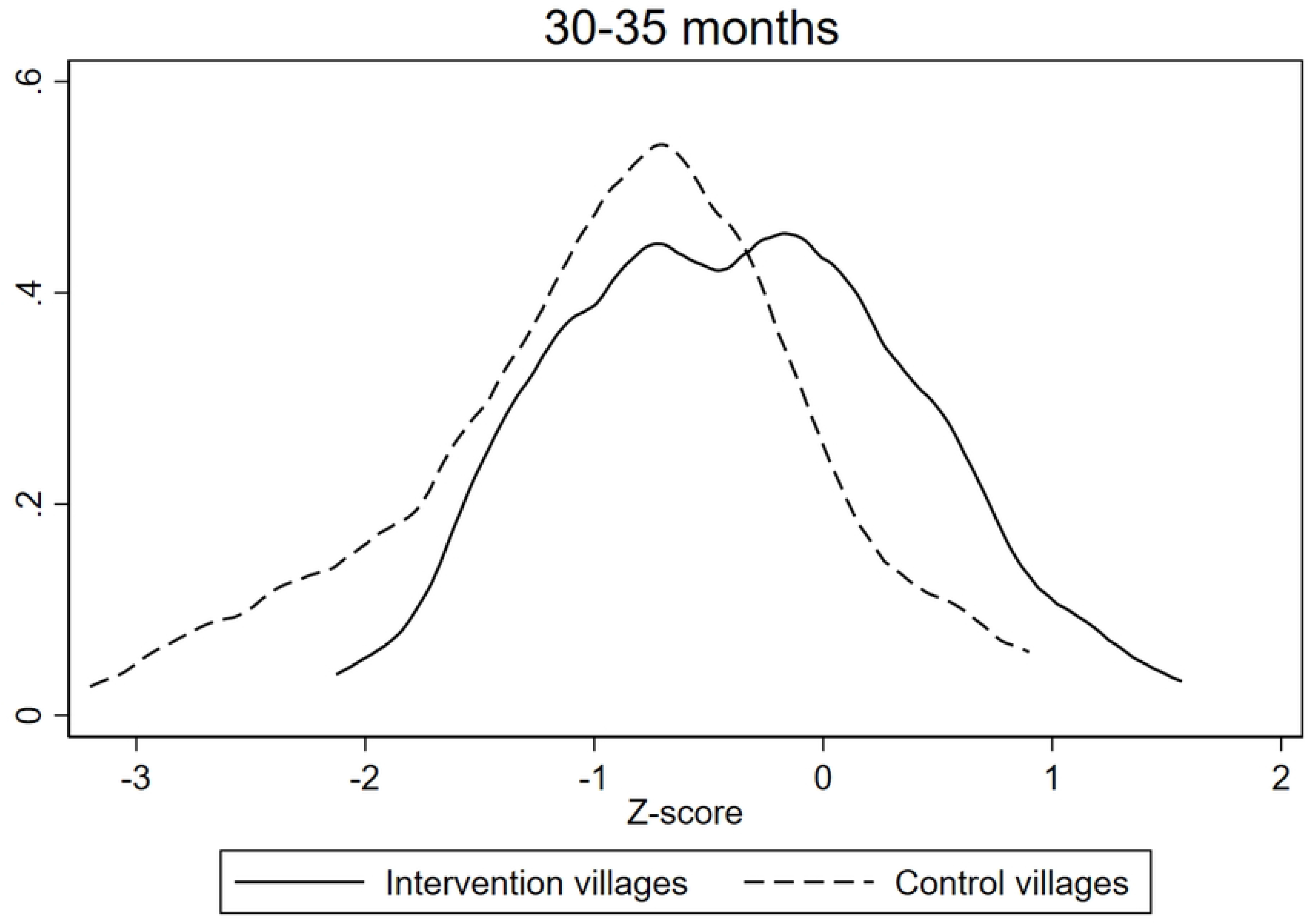
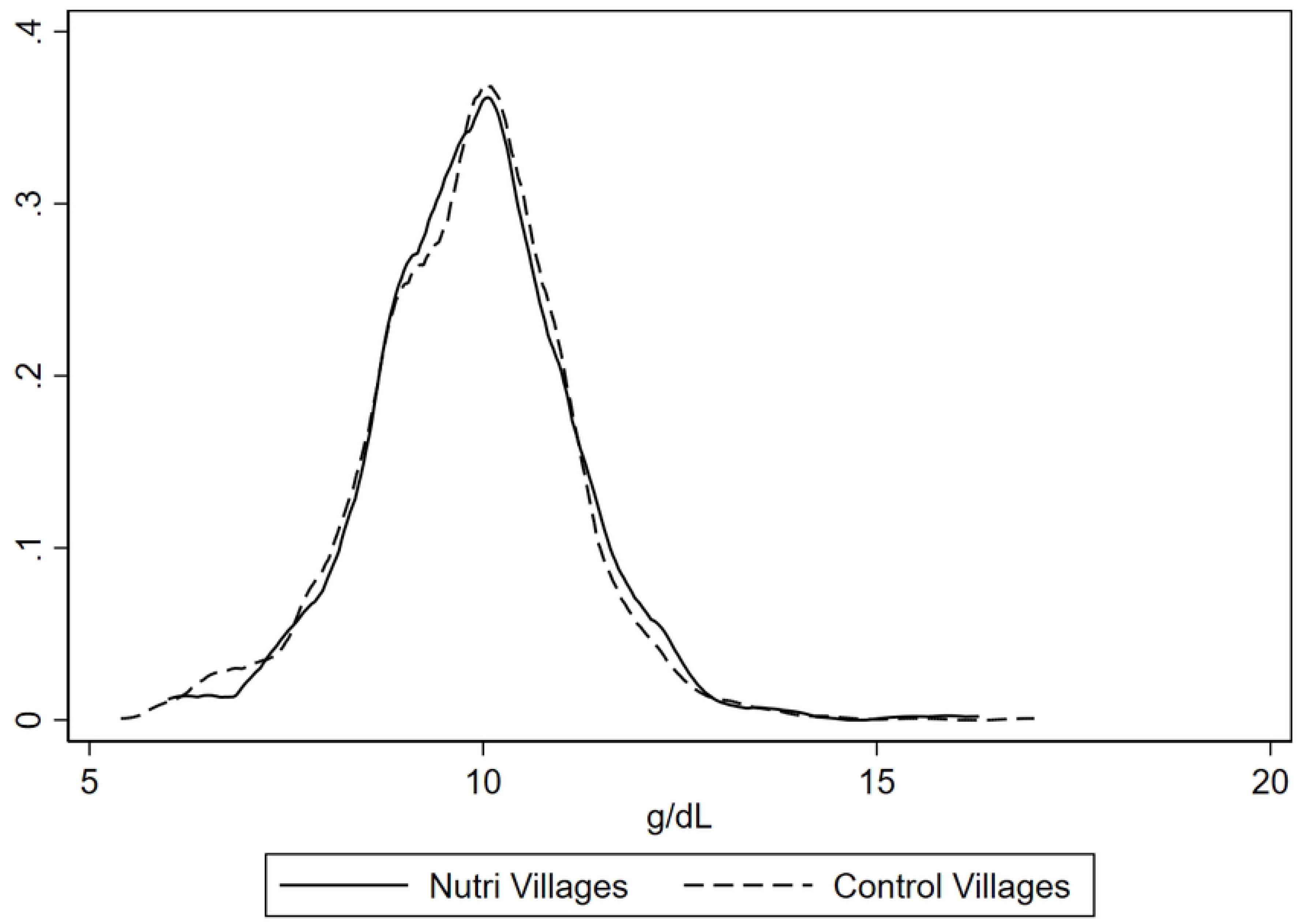
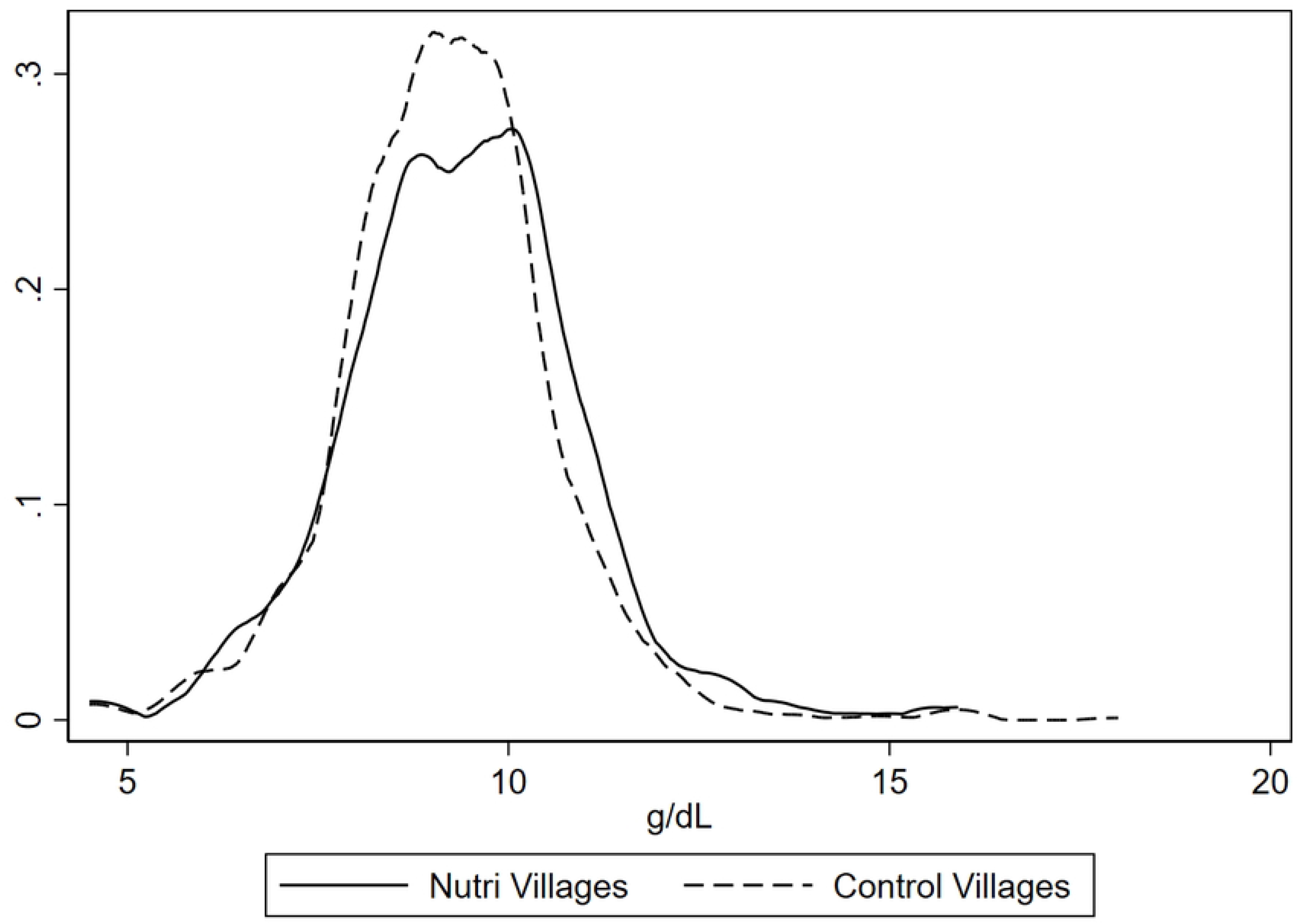
*A map depicting the 50 intervention villages and 79 control villages across the 13 districts of Andhra Pradesh (as of April 2022, there were 26 districts).*

### Evaluation Design and Sample

A cross-sectional survey was conducted between 20 December 2021 and 11 March 2022. Data were collected on dietary diversity for men, women, and children; nutritional status (anthropometry and anemia) for women and children, and child development for children under 3 years of age living in the 50 intervention villages and in 79 APCNF villages where the food-based nutrition intervention had not yet been implemented (herein ‘control villages’). The 79 control villages were villages adjacent to the intervention villages.

### Ethics

The Public Health Foundation of India Institutional Ethics Committee (Protocol number: TRC-IEC 488/22) and the Harvard T.H. Chan School of Public Health Institutional Review Board (Protocol number: IRB22-0220) approved the study. Written informed consent was obtained from all adult participants and parents/caregivers of children and adolescents.

### Data Collection

Data were collected by the NFFs, who were trained over two days in standardized data collection procedures and electronic data capture using EpiCollect. The survey questionnaire was pilot tested for suitability. NFFs collected the data in collaboration with the Department of Women and Child Development and local childcare centers known as Anganwadi Centers. The survey collected information on socio-economic and demographic characteristics, nutri-garden practices, and natural farming practices. LD, VJ, VC and SY had access to participant identifying information during data collection.

Dietary diversity for men, women, and children was assessed over the past 24 hours based on Food and Agriculture Organization guidelines (20). Women reported their own intake, and that of their children and spouse. A dietary diversity score (DDS, range 0-10) was calculated for women, men, and children over 2 years of age. A binary indicator was calculated for meeting minimum dietary diversity (MDD) if DDS was ≥5 food groups (20) . For children 6-24 months of age, DDS (range 0-8) and MDD (DDS≥5) were calculated according to the World Health Organization (WHO) Infant and Young Child Feeding (IYCF) indicators (21). Diet adequacy for breastfed children was calculated as children receiving ≥4 food groups and a minimum meal frequency (MMF), defined as receiving solid or semi-solid food ≥2 times per day for children 6-8 months of age and ≥3 times per day for children 9-23 months of age. Diet adequacy for non-breastfed children was calculated as meeting the following three IYCF practices: (1) being fed with other milk or milk products ≥2 times per day, (2) meeting MMF, defined as receiving solid or semi-solid foods ≥1 per day, and (3) receiving solid or semi-solid foods from ≥4 food groups not including the dairy food group (21).

Anthropometry for women and children under 2 years of age was measured using standardized procedures. Weight was measured to the nearest 0.1 gram using a digital or analogous scale. Height for women was measured to the nearest 0.1 cm using a stadiometer, and length for children under 2 years of age was measured using a length board. Women’s body mass index (BMI, kg/m^2^) was calculated, and women were categorized as underweight (BMI<18.5 kg/m^2^), normal weight (BMI between 18.5-24.9 kg/m^2^), overweight (BMI between 25-29.9 kg/m^2^), or obese (BMI≥30 kg/m^2^). Length-for-age Z-score (LAZ), weight-for-age Z-score (WAZ), and weight-for-length Z-score (WLZ) were calculated according to the 2006 WHO Child Growth Standards (22). Stunting was defined as LAZ <-2 SD, underweight as WAZ <-2 SD, and wasting as WLZ <-2 SD.

Hemoglobin (Hb) concentrations for women and children under 2 years of age were measured using a point-of-care device, *Mission*® Hemoglobin system. Among pregnant women and children under 2 years of age, anemia was defined as severe: Hb <7.0 g/dL, moderate: Hb 7-9.9 g/dL, mild: Hb 10-10.9 g/dL, and no anemia Hb ≥11 g/dL. Among non-pregnant women, anemia was defined as severe: Hb <8.0 g/dL, moderate: Hb 8-10.9 g/dL, mild: Hb 11-11.9 g/dL, and no anemia Hb ≥12 g/dL (23).

The short-form Caregiver Reported Early Development Index (CREDI) tool was used to assess child’s development in children under 3 years of age (24). The CREDI short form consists of a set of 20 items differing according to the following age bands: 0-5 months, 6-11 months, 12-17 months, 18-23 months, 24-29 months, and 30-35 months. The CREDI was administered by NFFs to the child’s primary caregiver who reported on whether the child can perform each item (yes/no) (25). Norm-referenced Z-scores for overall development for each age band were calculated from raw scores using the CREDI Scoring Application. A positive Z-score indicated a child has better developmental status than the average child in the CREDI reference population in the same age band. A Z-score of zero indicated a child has similar developmental status to the average child in the CREDI reference population in the same age band. A negative Z-score indicated a child has worse developmental status than the average child in the CREDI reference population in the same age band (26).

### Data Analysis

The analytic sample size included 3,511 households (1,121 intervention households and 2,390 control households). We calculated mean and SD for continuous variables and proportions for binary variables. We used t-tests to test whether differences between the control and the intervention groups were significant. P-value <0.05 was considered statistically significant. Data analysis was conducted using Stata 17.0.

## Results

Sample characteristics are summarized in Table 1. Women were, on average, 25 years old. Forty-three percent of intervention villages were classified as ‘tribal villages’ versus 23% of control villages (p<0.01). Villages were classified as ‘tribal’ based on standard Government of India criterion, e.g., more than 25% of the village population belonged to Scheduled Tribe (27). Thirty-seven percent of the households in intervention villages reported practicing natural farming compared to 12% in control villages (p<0.01). Forty-three percent of households in intervention villages reported having a nutri-garden compared to 11% of households in control villages (p<0.01). Thirty-nine percent of households in intervention villages reported having backyard poultry compared to 21% in control villages (p<0.01). Of those who had backyard poultry, 8% of households in intervention villages reported selling and not consuming their backyard poultry produce compared to 18% in control villages – in other words, households in control villages were more likely to sell their produce and not consume them than households in intervention villages (p<0.01).

**Table 1.**
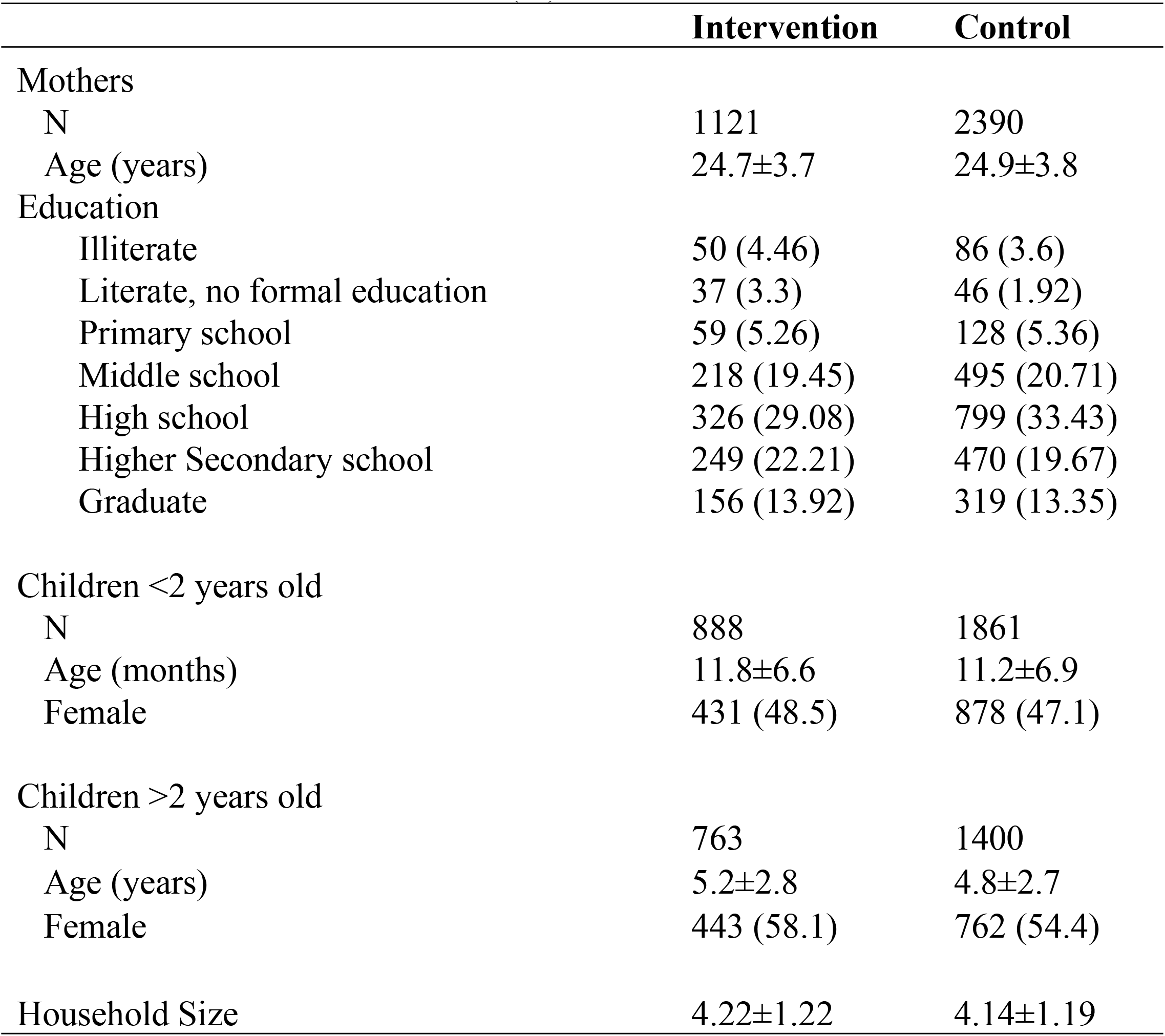
Descriptive characteristics of the 3,511 households enrolled in the evaluation of a nutrition-sensitive agroecology program in Andhra Pradesh, India. Values are Mean ± SD or N (%).

|  | Intervention | Control |
| --- | --- | --- |
| Mothers |  |  |
| N | 1121 | 2390 |
| Age (years) | 24.7±3.7 | 24.9±3.8 |
| Education |  |  |
| Illiterate | 50 (4.46) | 86 (3.6) |
| Literate, no formal education | 37 (3.3) | 46 (1.92) |
| Primary school | 59 (5.26) | 128 (5.36) |
| Middle school | 218 (19.45) | 495 (20.71) |
| High school | 326 (29.08) | 799 (33.43) |
| Higher Secondary school | 249 (22.21) | 470 (19.67) |
| Graduate | 156 (13.92) | 319 (13.35) |
| Children <2 years old |  |  |
| N | 888 | 1861 |
| Age (months) | 11.8±6.6 | 11.2±6.9 |
| Female | 431 (48.5) | 878 (47.1) |
| Children >2 years old |  |  |
| N | 763 | 1400 |
| Age (years) | 5.2±2.8 | 4.8±2.7 |
| Female | 443 (58.1) | 762 (54.4) |
| Household Size | 4.22±1.22 | 4.14±1.19 |

### Dietary Diversity

DDS among women and men was mean (SD) 6.53 (±1.62) and 6.16 (±1.65), respectively in intervention villages and 5.81 (±1.58) and 5.39 (±1.61), respectively in control villages (Supplementary Table T1). Adults in intervention villages were more likely to have diverse diets compared to adults in control villages: 87% of women and 83% of men in intervention villages had a diverse diet (DDS ≥5) compared to 79% of women and 73% of men in control villages (p<0.01 for women and p<0.01 for men). Compared to women in control villages, women in intervention villages were significantly more likely to consume all food groups *except* starchy staples, other vegetables, and flesh foods (p<0.01) (Fig 2, panel A). Results were similar for men (Fig 2, panel B).

**Figure 2.**
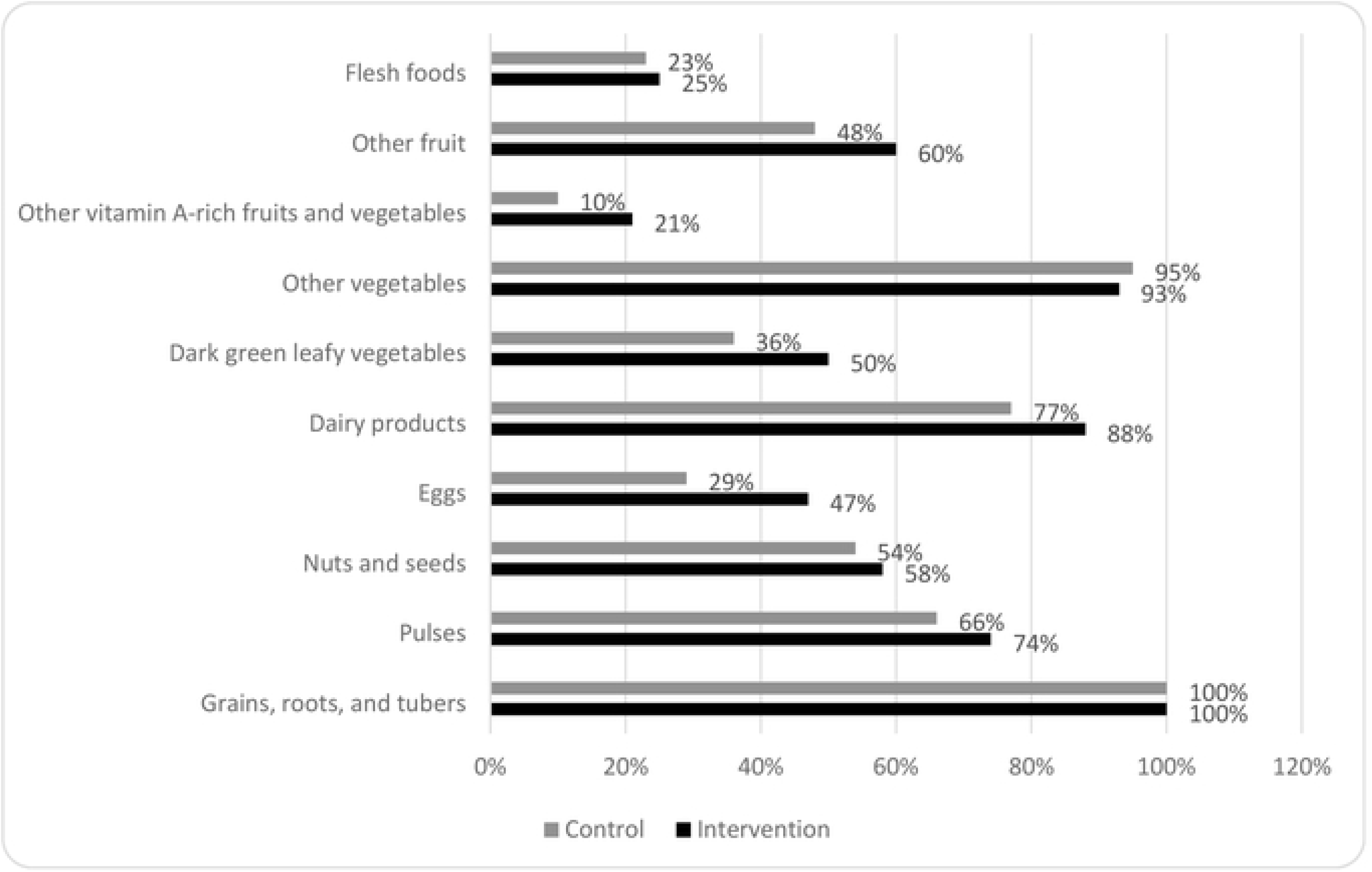
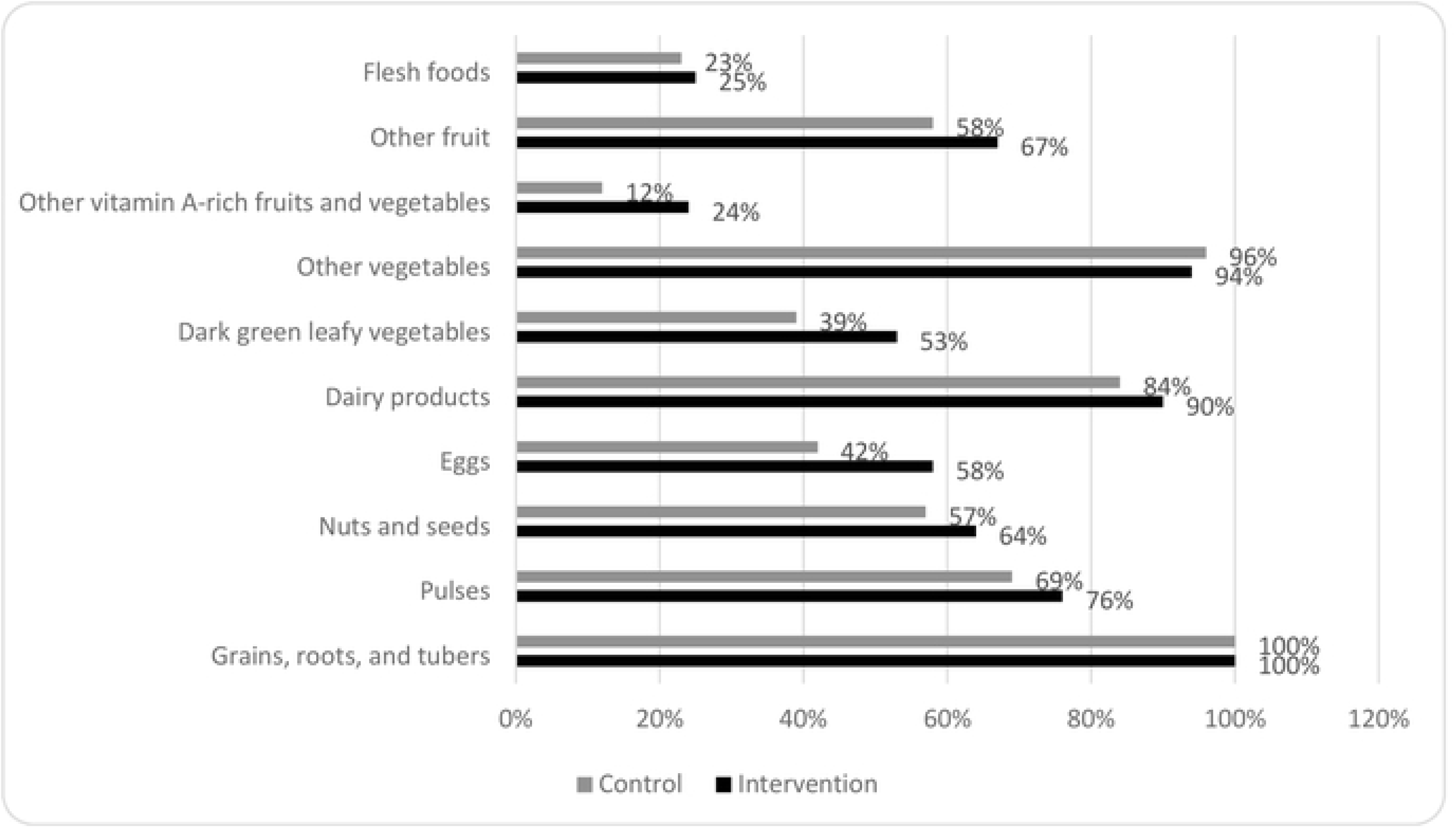
*Food groups consumed by (A) adult women and (B) adult men in intervention versus control villages as part of an evaluation of a nutrition-sensitive agroecology program in Andhra Pradesh, India. P<0.01 for all intervention versus control comparisons except starchy staples, other vegetables, and flesh foods.*

Forty-three percent of women and 41% of men in intervention villages reported consuming any type of naturally farmed foods compared to 22% and 15%, respectively, in control villages. Overall, consumption of naturally farmed starchy staples, pulses, dark green leafy vegetables, vitamin A-rich fruits and vegetables, and other fruits and vegetables, was higher in intervention villages as compared to control villages (Supplementary Fig S1).

In children 6-24 months of age, 8% of breastfed children in intervention villages had an adequate diet compared to 11% in control villages (p=0.02). DDS among children 6-24 months of age in intervention and control villages was mean (SD) 2.99 (±1.52) and 2.73 (±1.62), respectively (p<0.01) (Supplementary Table T2). With regards to specific food groups, children 6-24 months of age in intervention villages were more likely to consume eggs and vitamin A-rich fruits and vegetables (Fig 3, panel A). No differences were observed for the other food groups. There were no differences in DDS by child sex (p=0.12).

**Figure 3.**
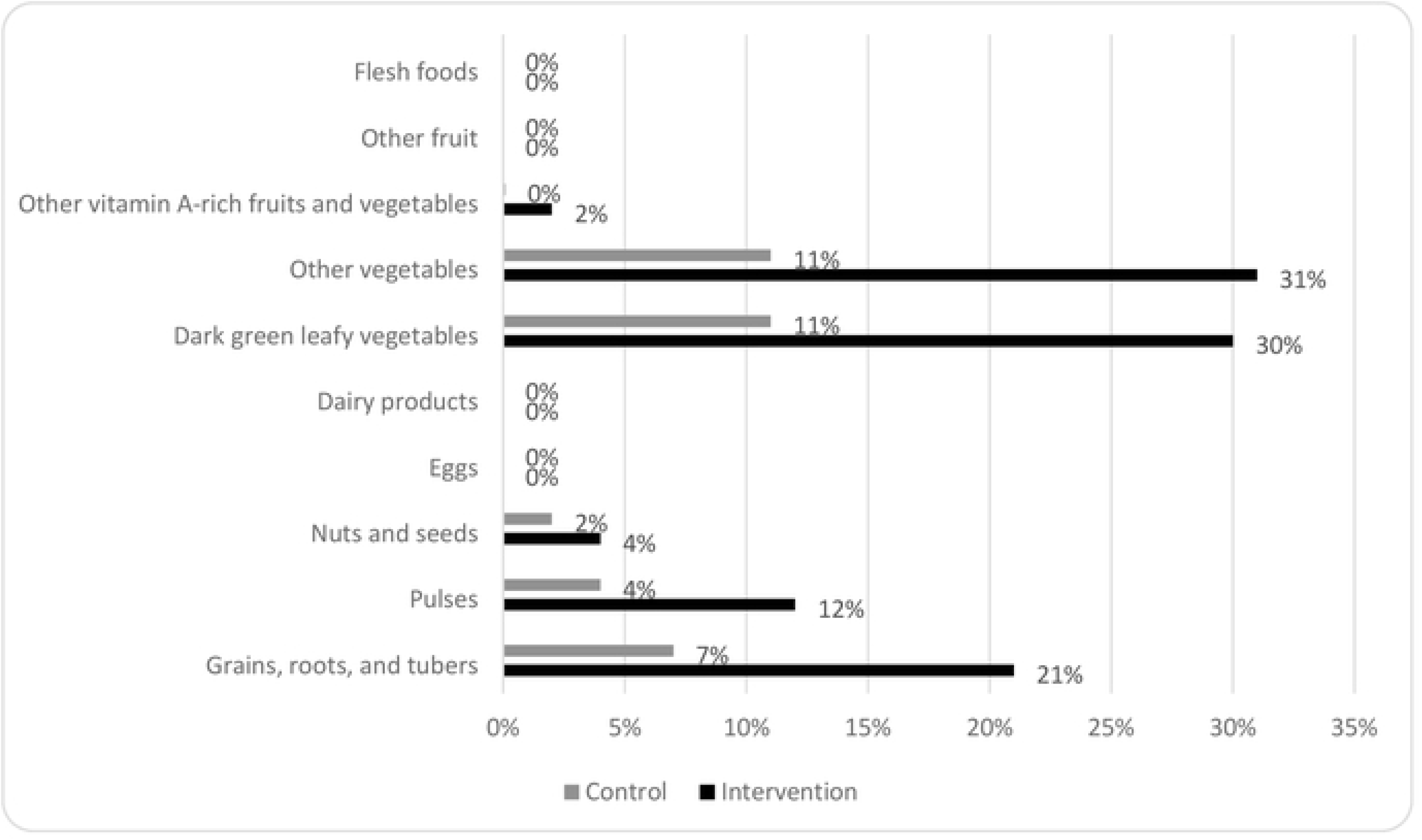
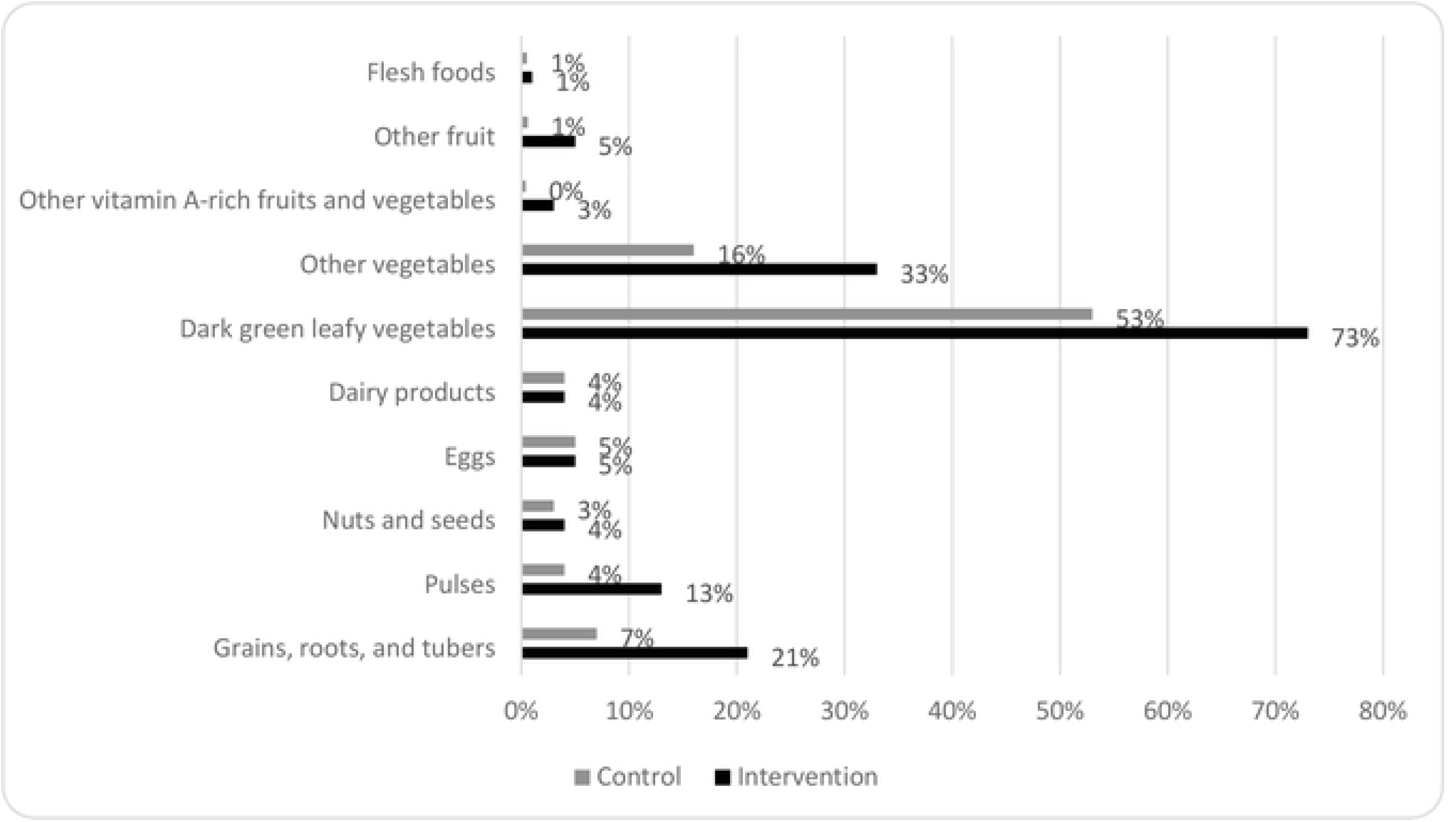
*Food groups consumed by (A) children under 2 years old and (B) children over 2 years old in intervention versus control villages as part of an evaluation of a nutrition-sensitive agroecology program in Andhra Pradesh, India.*

Among children over 2 years of age (range 2-18 years), DDS was mean (SD) 6.6 (±1.7) in intervention villages and 6.0 (±1.5) in control villages (p<0.01). In the intervention villages, children over 2 years of age were more likely to consume other fruits, vitamin A-rich fruits and vegetables, dark leafy vegetables, dairy products, nuts, and eggs (Fig 3, panel B). There were no differences in DDS by child sex in children over 2 years of age (p=0.16).

### Nutritional Status

Women in intervention villages were less likely to be overweight (16% versus 23%, respectively p=0.01) and more likely to be underweight as compared to women in control villages (12% versus 8%, respectively, p=0.02) (Fig 4, panel A). In children under 2 years of age, stunting prevalence was similar in intervention and control villages (p=0.83) (Fig 4, Panel A). However, children in intervention villages were more likely to be underweight (28% versus 23%, respectively, p=0.01) and wasted (26% versus 21%, respectively, p<0.01) compared to children in control villages. Of note, children measured with an analogue scale were twice as likely to be wasted as children measured with a digital scale (23% versus 11%, respectively, p<0.01), whereas women measured with an analogue scale were less likely to be underweight than those measured with a digital scale (9% versus 17%, respectively, p=0.02).

**Figure 4.**
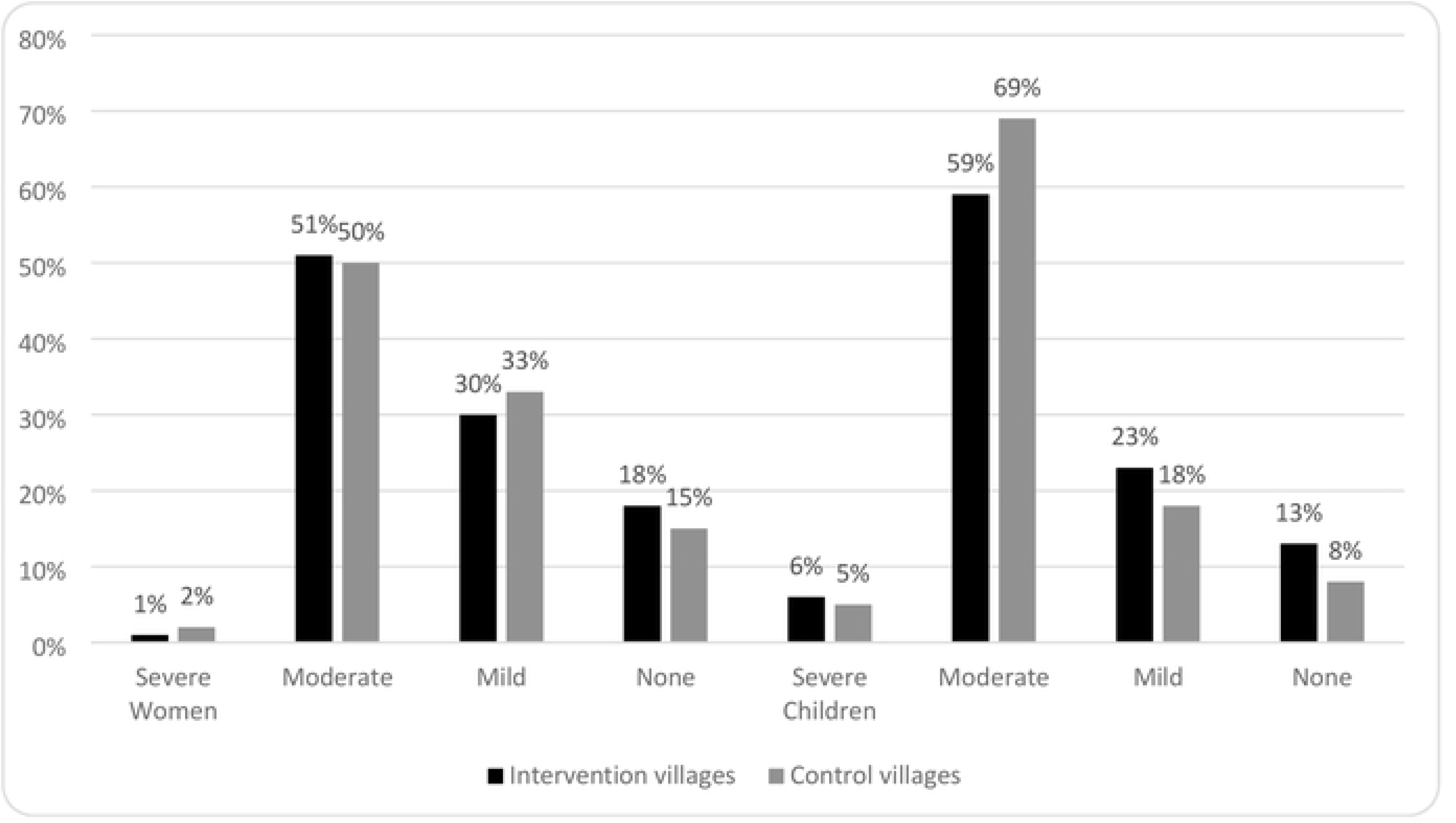
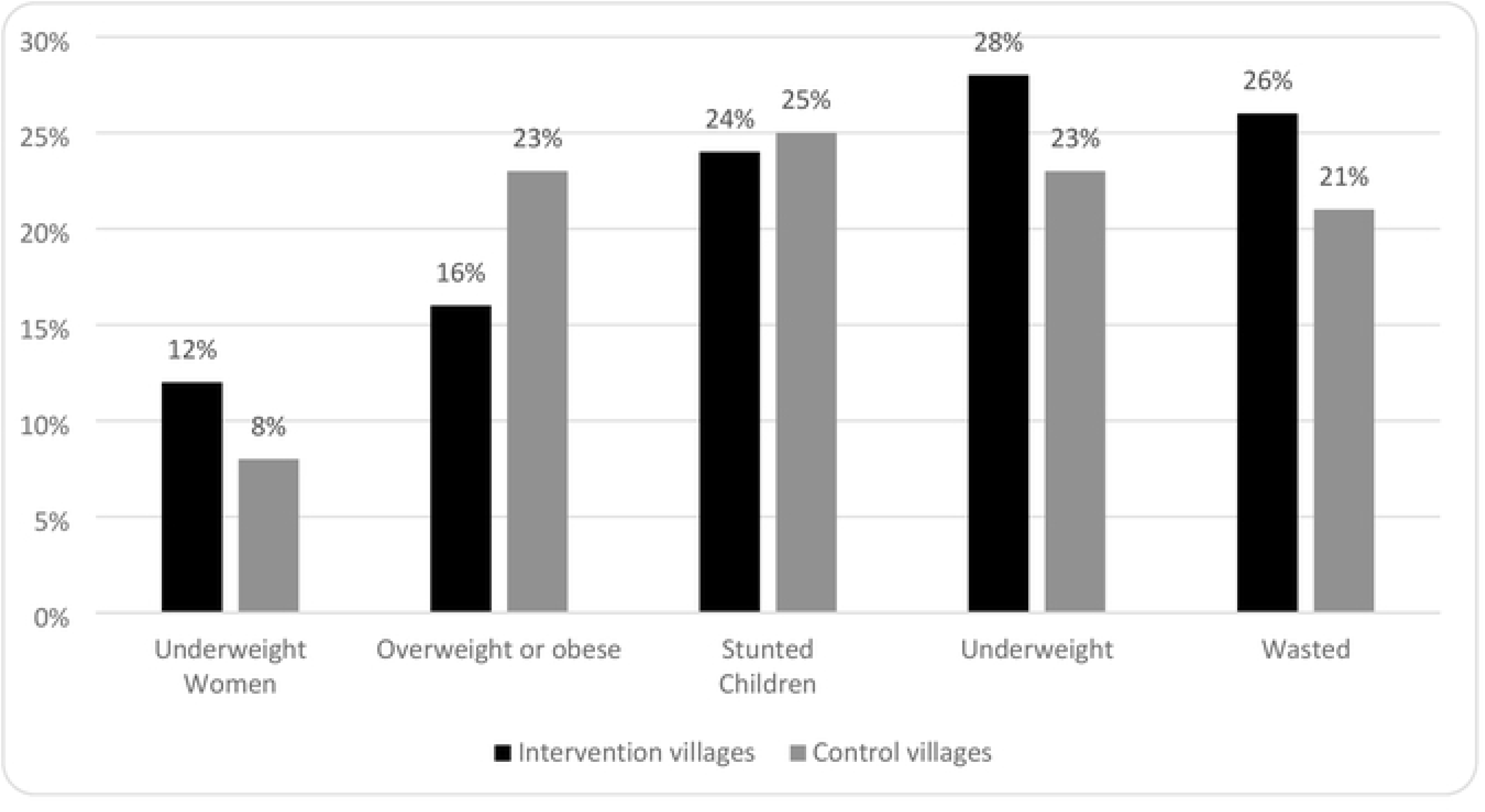
*Nutritional status of women and children under 2 years of age, including (A) anthropometry and (B) anemia in intervention and control villages as part of an evaluation of a nutrition-sensitive agroecology program in Andhra Pradesh, India.*

Moderate anemia was the most common form of anemia, with half of women in both intervention and control villages classified as having moderate anemia (Fig 4, panel B). There were no significant differences in anemia prevalence between women in intervention versus control villages. In contrast, children under 2 years of age in intervention villages were less likely to be anemic as compared to control villages: 59% versus 69%, respectively (p<0.01) (Fig 4, panel B). When examining the distribution of hemoglobin concentrations, the entire distribution had shifted right in intervention villages, i.e., to higher hemoglobin values among children (Supplementary Fig S2).

### Child Development

Children in intervention villages had better early child development scores as compared to children in control villages with larger benefits among older age groups (Fig 5). We observed a shift from negative Z-score to positive Z-scores with increase in child age. Among children 0-5 months of age, children in control villages had better Z-scores than children in intervention villages: -0.37 versus -0.68, respectively (p<0.01). Among children 6-11 months and 12-17 months old, children in intervention villages had consistently higher Z-scores, albeit not statistically significant (Supplementary Table T3). However, among children 18-23 months of age, those in intervention villages had significantly higher mean Z-scores compared to children in control villages (0.46 versus 0.24, respectively, p=0.02). Among children 24-29 months of age, those in intervention villages had significantly higher Z-scores compared to children in control villages (-0.14 versus -0.53 respectively, p<0.01). Among children 30-35 months of age, those in intervention villages had significantly higher Z-scores relative to children in control villages (-0.38 versus -0.91 respectively, p<0.01).

**Figure 5.**
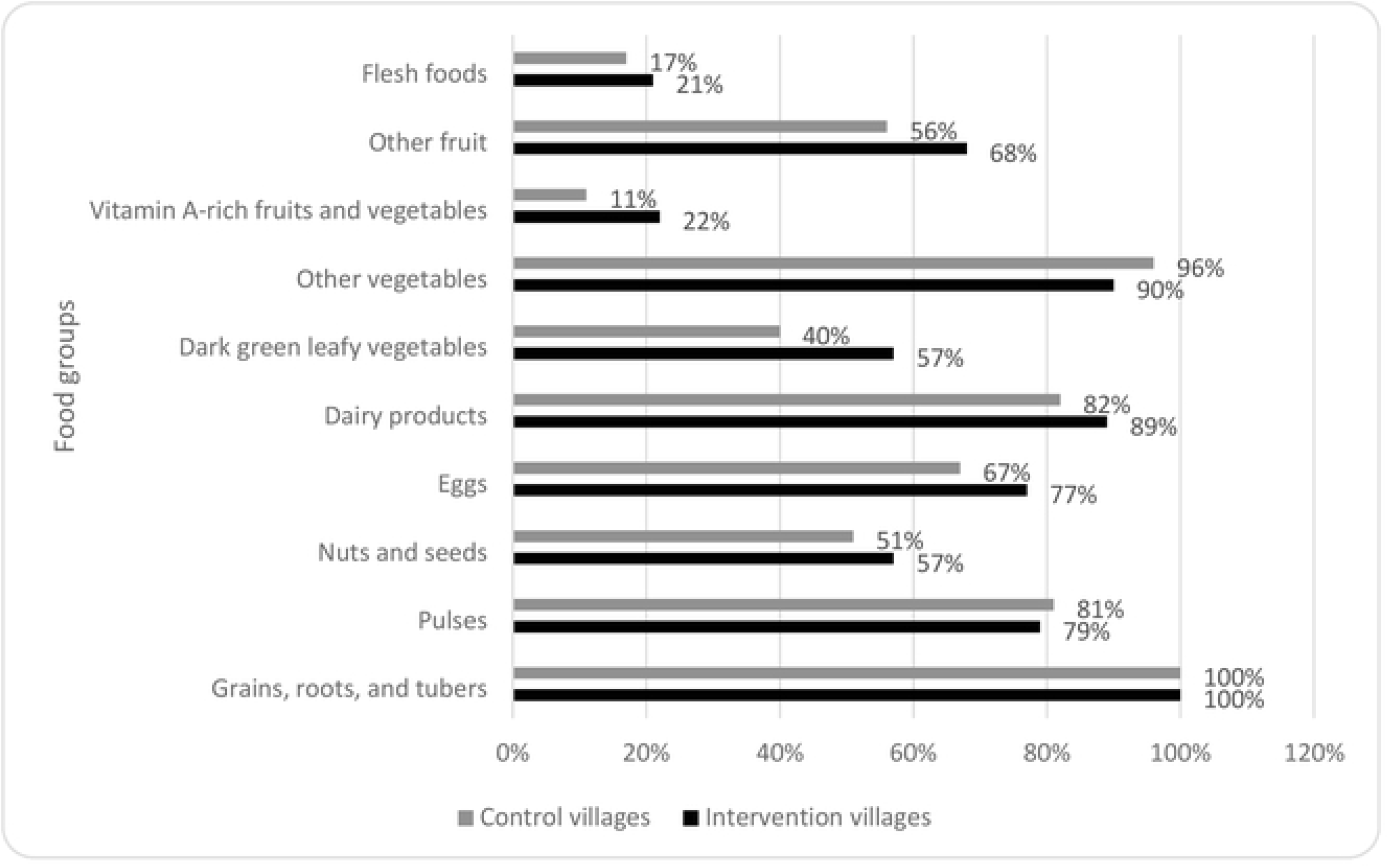
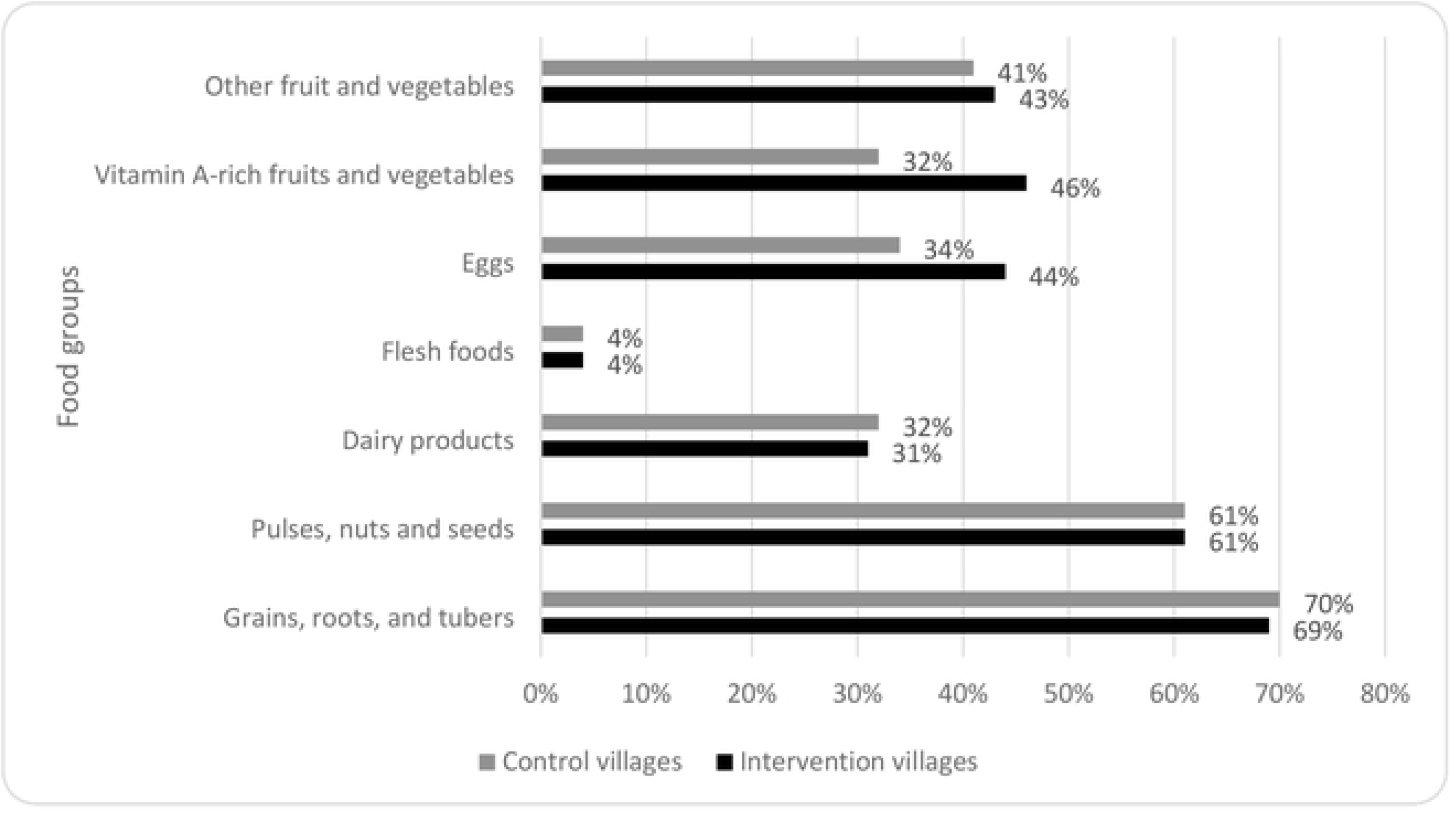
*Norm-referenced child development Z-scores for children under 3 years of age in intervention and control villages as part of an evaluation of a nutrition-sensitive agroecology program in Andhra Pradesh, India.*

## Discussion

In this cross-sectional program evaluation, we explored differences in dietary diversity, nutritional status, and child development between adults and children in villages receiving a nutrition-sensitive agroecological intervention and control villages where only an agroecological intervention was implemented. We found that women, children, and men in intervention villages had more diverse diets and consumed more naturally farmed foods relative to those in control villages. Children under 2 years of age also had improved hemoglobin status. Child development scores were also higher in intervention villages compared to control, particularly among older children 24-36 months of age. These findings suggest that the additional nutrition-sensitive activities implemented in intervention villages were effective in improving diets, nutrition, and child development.

The findings from our evaluation show that the intervention villages tended to have more diverse diets compared to control villages. These values are slightly higher (indicating slightly better feeding practices) than NFHS-5 for rural Andhra Pradesh (6% of breastfed children had an adequate diet versus 11% in our evaluation). The higher consumption of eggs and green leafy vegetables in the intervention villages may be attributed to various nutrition-sensitive initiatives undertaken as part of the intervention such as homestead food production including backyard poultry and nutri-gardens. The findings from this evaluation align with the results obtained in other studies conducted in India, where planting of nutri gardens was also associated with increased consumption of eggs and green leafy vegetables (10,11).

Consumption of eggs and vitamin A-rich fruit and vegetables was higher in children 6-24 months age in intervention villages. Similarly, children older than 2 years of age consumed more eggs, vitamin A-rich fruits and vegetables, dark green leafy vegetables, dairy products, other fruits, flesh foods and nuts compared to their counterparts in the control villages. These findings support the effectiveness of nutri-gardens and BCC activities aimed at influencing the feeding practices in increasing the consumption of diverse foods.

Despite having more diverse diets, children under 2 years of age and mothers in intervention villages were more likely to be underweight than those in control villages. The findings may be partly explained by the fact that intervention villages were more likely to be tribal villages than control villages, and tribal villages historically have higher levels of underweight (28). Another explanation for this counter-intuitive finding could be differences in measurement instruments used in intervention versus control villages with NFFs in control villages being more likely to use an analogue scale as compared to NFFs in intervention villages (98% versus 91%, respectively, p<0.01). The scales used reflected the scales available at Anganwadi Centers as these were used for the evaluation.

Despite the nutrition-sensitive agroecology program, moderate and mild anemia were common in intervention and control villages. This could be partly explained by the complex etiology of anemia – it is not just diet that contributes. In addition, water and sanitation access and behaviors could influence the absorption of nutrients, which in turn could influence anemia (29). In addition, the high prevalence of anemia in children under 2 years of age in our sample (90%) could be due to the instrument used to measure hemoglobin. For example, NFHS-5, which uses capillary blood, estimated a prevalence of anemia among children under 5 years of age in Andhra Pradesh of 63% (17). In contrast, the Comprehensive National Nutrition Survey (CNNS, 2016-18), which uses venous blood, estimated a prevalence in the same age group of 40% (30). The Mission® Hb device used in this evaluation is a point-of-care device which uses capillary blood. A study in Malaysia showed that the Mission® Plus Hb device underestimated hemoglobin relative to the gold standard cyanmethemoglobin method (31), which would result in an over-estimation of anemia prevalence.

Nonetheless, both intervention and control villages in our evaluation used the same device and so this measurement error was not differential between intervention and control villages. We observed that while there were no statistically significant differences in women’s anemia status, children in intervention villages were less likely to be anemic than children in control villages. This is especially notable considering the fact that intervention villages were more likely to be tribal villages, where the baseline anemia prevalence tends to be higher (32). This improved anemia status in children could be explained by the improved diets among children in intervention villages.

We also found large, consistent, and beneficial effects on child development, with larger benefits among older children 24-36 months of age. There are several plausible explanations for this. First, children in intervention villages may have had lower pesticide exposure due to eating naturally farmed foods. Organophosphorus pesticides, which are commonly used in Andhra Pradesh(33), are known acetylcholinesterase inhibitors contributing to neurocognitive impairment in children (15). Second, children in intervention villages were slightly less likely to have anemia, a known risk factor for suboptimal child development (34). Together with improved diet and better dietary diversity scores, this may have improved their energy levels and thus their engagement with caregivers and ultimately their development.

Our results from a single time point of a non-randomized intervention should be interpreted with caution. To estimate the combined effects of natural farming with and without nutrition-sensitive activities, a randomized control trial should be conducted to establish the causal relationship. Co-Benefits of Large-Scale Organic FarMing on HuMan Health (BLOOM) is an upcoming cluster-randomized evaluation looking at the effectiveness of APCNF on urinary pesticides and dietary diversity (35). Further, randomized evaluations are needed to assess the additional benefits of nutrition-sensitive activities and to help disentangle which component of the nutrition-sensitive package worked (homestead food production, BCC, nutrition counselling, or cooking demonstrations) or whether it was the package as a whole.

Additional limitations include women reporting the dietary diversity of men and children (particularly for older children who may eat a lot outside the home) (36), which could have led to reporting bias. Moreover, the reported dietary data does not include information on quantities or micronutrient intake, which would help us better understand the deficiencies and the possible solutions to tackle them. Also, the women may have over-reported their consumption of healthy foods emphasized during BCC sessions since the data collectors (the NFFs) were also the implementers, leading to a social-desirability bias. As mentioned above, there were also differences in anthropometric measures by use of different scales (analogue vs digital). Future studies should use digital scales to measure weight and venous blood samples to measure anemia to get more accurate estimates.

## Conclusion

In conclusion, this nutrition-sensitive agroecological program improved women, men, and child dietary diversity, reduced anemia in children under 2 years of age, and improved child development in children under 3 years of age. These findings suggest that the nutrition-sensitive activities (homestead food production, BCC, cooking demonstration, and nutrition counselling) were effective. Effects on child development were likely also achieved through the increased consumption of naturally farmed foods. The policy direction provided by the Ministry of Women and Child Development, Government of India, in establishing Poshan Vatikas (nutri-gardens) at Anganwadi Centers under Mission POSHAN could benefit from integrating agroecological principles. Integrating nutrition-sensitive interventions with agroecological interventions can be more effective and aid the achievement of ‘Anemia mukt Bharat’ (Anemia Free India). These initiatives will not only enhance food availability, accessibility, and consumption at the household level, but also contribute to safeguarding against the adverse effects of pesticides. The Government of India’s commitment to agroecological practices (i.e., natural farming) was recently reaffirmed in the 2023-24 budget speech which announced that over the next three years, the government will facilitate 10 million farmers to adopt these practices (37). Our evaluation of a nutrition-sensitive agroecological program in Andhra Pradesh suggests that better outcomes can be achieved if synergies are leveraged between various departments towards achieving holistic nutrition and development.

## Data Availability

The data underlying the results presented in the study are available upon reasonable request to Lakshmi Durga.

## Acknowledgements

We would like to thank the Nutrition Farming Fellows who collected the data for this evaluation, and the participants for kindly volunteering their time for assessments. We would also like to thank Govinda Raju for preparing the map of participating villages.

## Supplementary Material

**Supplementary Table T1:**
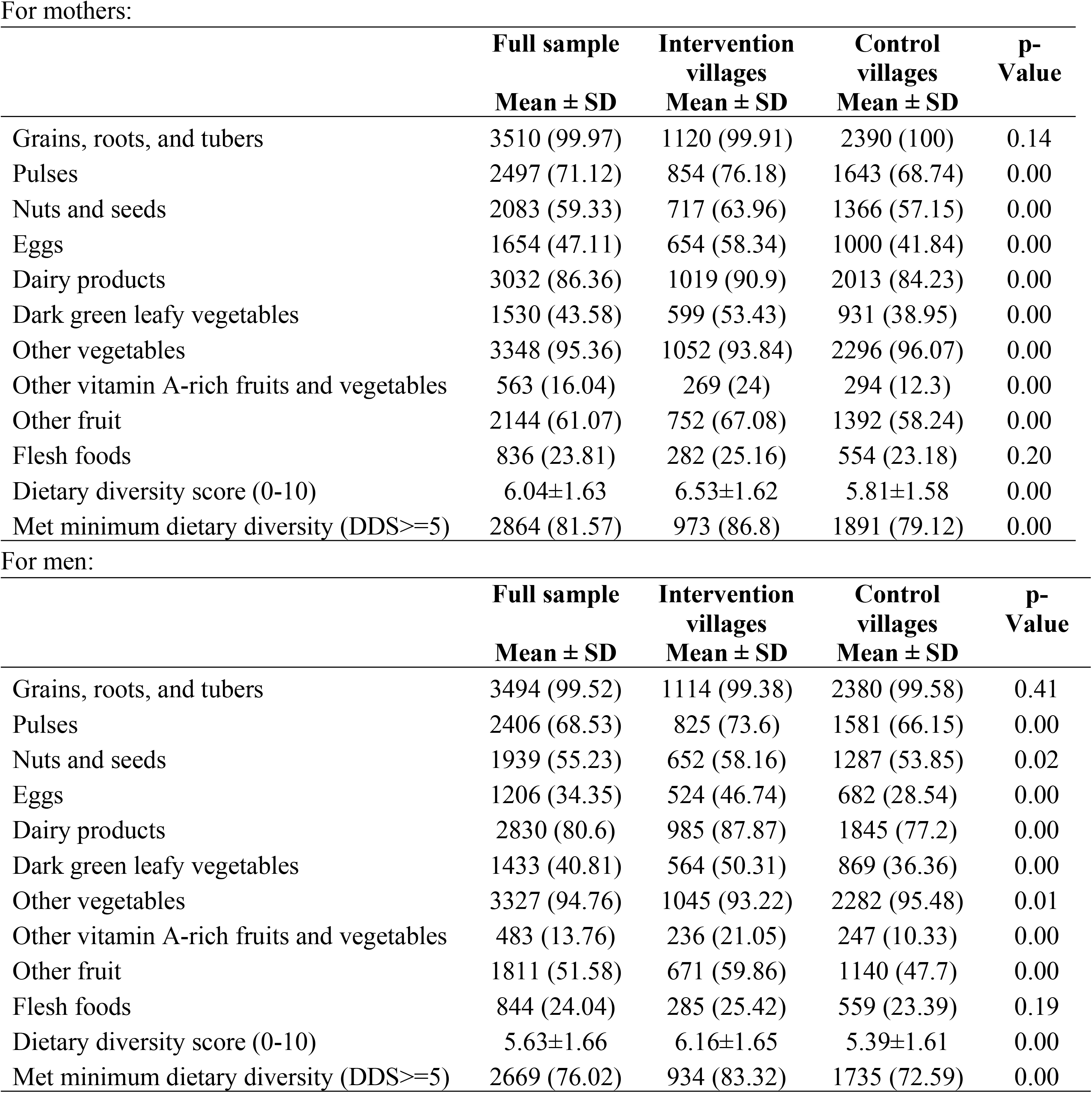
Dietary Diversity Scores among men and women

|  | **Full sample** | **Intervention villages** | **Control villages** | **p-Value** |
| --- | --- | --- | --- | --- |
|  | **Mean ± SD** | **Mean ± SD** | **Mean ± SD** |  |
| Grains, roots, and tubers | 3510 (99.97) | 1120 (99.91) | 2390 (100) | 0.14 |
| Pulses | 2497 (71.12) | 854 (76.18) | 1643 (68.74) | 0.00 |
| Nuts and seeds | 2083 (59.33) | 717 (63.96) | 1366 (57.15) | 0.00 |
| Eggs | 1654 (47.11) | 654 (58.34) | 1000 (41.84) | 0.00 |
| Dairy products | 3032 (86.36) | 1019 (90.9) | 2013 (84.23) | 0.00 |
| Dark green leafy vegetables | 1530 (43.58) | 599 (53.43) | 931 (38.95) | 0.00 |
| Other vegetables | 3348 (95.36) | 1052 (93.84) | 2296 (96.07) | 0.00 |
| Other vitamin A-rich fruits and vegetables | 563 (16.04) | 269 (24) | 294 (12.3) | 0.00 |
| Other fruit | 2144 (61.07) | 752 (67.08) | 1392 (58.24) | 0.00 |
| Flesh foods | 836 (23.81) | 282 (25.16) | 554 (23.18) | 0.20 |
| Dietary diversity score (0-10) | 6.04±1.63 | 6.53±1.62 | 5.81±1.58 | 0.00 |
| Met minimum dietary diversity (DDS≥5) | 2864 (81.57) | 973 (86.8) | 1891 (79.12) | 0.00 |

For men:
|  | **Full sample** | **Intervention villages** | **Control villages** | **p-Value** |
| --- | --- | --- | --- | --- |
|  | **Mean ± SD** | **Mean ± SD** | **Mean ± SD** |  |
| Grains, roots, and tubers | 3494 (99.52) | 1114 (99.38) | 2380 (99.58) | 0.41 |
| Pulses | 2406 (68.53) | 825 (73.6) | 1581 (66.15) | 0.00 |
| Nuts and seeds | 1939 (55.23) | 652 (58.16) | 1287 (53.85) | 0.02 |
| Eggs | 1206 (34.35) | 524 (46.74) | 682 (28.54) | 0.00 |
| Dairy products | 2830 (80.6) | 985 (87.87) | 1845 (77.2) | 0.00 |
| Dark green leafy vegetables | 1433 (40.81) | 564 (50.31) | 869 (36.36) | 0.00 |
| Other vegetables | 3327 (94.76) | 1045 (93.22) | 2282 (95.48) | 0.01 |
| Other vitamin A-rich fruits and vegetables | 483 (13.76) | 236 (21.05) | 247 (10.33) | 0.00 |
| Other fruit | 1811 (51.58) | 671 (59.86) | 1140 (47.7) | 0.00 |
| Flesh foods | 844 (24.04) | 285 (25.42) | 559 (23.39) | 0.19 |
| Dietary diversity score (0-10) | 5.63±1.66 | 6.16±1.65 | 5.39±1.61 | 0.00 |
| Met minimum dietary diversity (DDS≥5) | 2669 (76.02) | 934 (83.32) | 1735 (72.59) | 0.00 |

**Supplementary Table T2:** Dietary Diversity Scores among Children <2 and >2 years age For Children 6-23 months age:

|  | **Full sample** | **Intervention villages** | **Control villages** | **p-value** |
| --- | --- | --- | --- | --- |
|  | **Mean ± SD** | **Mean ± SD** | **Mean ± SD** |  |
| Grains, roots, and tubers | 1410 (69.87) | 474 (69.1) | 936 (70.27) | 0.59 |
| Pulses, nuts, and seeds | 1232 (61.14) | 423 (61.66) | 809 (60.87) | 0.73 |
| Dairy products | 647 (31.98) | 216 (31.44) | 431 (32.26) | 0.71 |
| Flesh foods | 78 (3.87) | 29 (4.23) | 49 (3.68) | 0.54 |
| Eggs | 762 (37.67) | 305 (44.4) | 457 (34.21) | 0.00 |
| Vit- A rich fruits and vegetables | 734 (36.39) | 313 (45.69) | 421 (31.61) | 0.00 |
| Other fruits and vegetables | 837 (41.39) | 294 (42.86) | 543 (40.64) | 0.34 |
| Dietary diversity score (0-8) | 3.55±1.54 | 3.71±1.48 | 3.47±1.56 | 0.00 |
| Met minimum dietary diversity (DDS≥5) | 542 (26.79) | 207 (30.13) | 335 (25.07) | 0.02 |

For Children >2 years age:
|  | **Full sample** | **Intervention villages** | **Control villages** | **p-values** |
| --- | --- | --- | --- | --- |
|  | **Mean ± SD** | **Mean ± SD** | **Mean ± SD** |  |
| Grains, roots, and tubers | 2164 (99.95) | 763 (99.87) | 1401 (100) | 0.18 |
| Pulses | 1740 (80.37) | 603 (78.93) | 1137 (81.16) | 0.21 |
| Nuts and seeds | 1153 (53.26) | 434 (56.81) | 719 (51.32) | 0.01 |
| Eggs | 1526 (70.48) | 588 (76.96) | 938 (66.95) | 0.00 |
| Dairy products | 1829 (84.48) | 681 (89.14) | 1148 (81.94) | 0.00 |
| Dark green leafy vegetables | 996 (46) | 434 (56.81) | 562 (40.11) | 0.00 |
| Other vegetables | 2023 (93.44) | 685 (89.66) | 1338 (95.5) | 0.00 |
| Other vitamin A-rich fruits and vegetables | 319 (14.73) | 165 (21.6) | 154 (10.99) | 0.00 |
| Other fruit | 1312 (60.6) | 523 (68.46) | 789 (56.32) | 0.00 |
| Flesh foods | 393 (18.15) | 158 (20.68) | 235 (16.77) | 0.02 |
| Dietary diversity score (0-10) | 6.21±1.61 | 6.59±1.69 | 6.01±1.53 | 0.00 |
| Met minimum dietary diversity (DDS≥5) | 1832 (84.62) | 660 (86.39) | 1172 (83.65) | 0.09 |

**Supplementary Table T3:** Child Development (Z - Scores) among various age groups

|  | **Full sample** | **Intervention**<br>**villages** | **Control**<br>**villages** | **p-Value** |
| --- | --- | --- | --- | --- |
|  | **Mean ± SD** | **Mean ± SD** | **Mean ± SD** |  |
| *Child age: 0-5 months* |  |  |  |  |
| N | 2910 | 987 | 1923 |  |
| OVERALL | 47.26±4.6 | 47.79±4.6 | 46.99±4.58 | 0.00 |
| Z_OVERALL | -0.25±1.13 | -0.18±1.19 | -0.28±1.09 | 0.03 |
| *Child age: 6-11 months* |  |  |  |  |
| N | 662 | 221 | 441 |  |
| OVERALL | 45.03±1.84 | 45.23±1.95 | 44.92±1.78 | 0.04 |
| Z_OVERALL | -0.4±1.23 | -0.35±1.38 | -0.42±1.15 | 0.46 |
| *Child age: 12-17 months* |  |  |  |  |
| N | 646 | 235 | 411 |  |
| OVERALL | 49.09±1.29 | 49.05±1.36 | 49.12±1.25 | 0.48 |
| Z_OVERALL | -0.19±1.06 | -0.1±1.14 | -0.24±1.01 | 0.12 |
| *Child age: 18-23 months* |  |  |  |  |
| N | 538 | 185 | 353 |  |
| OVERALL | 51.58±1.16 | 51.67±1.24 | 51.53±1.12 | 0.16 |
| Z_OVERALL | 0.31±1.09 | 0.46±1.13 | 0.24±1.06 | 0.02 |
| *Child age: 24-29 months* |  |  |  |  |
| N | 239 | 82 | 157 |  |
| OVERALL | 52.05±0.88 | 52.3±0.94 | 51.92±0.82 | 0.00 |
| Z_OVERALL | -0.4±0.79 | -0.14±0.8 | -0.53±0.76 | 0.00 |
| *Child age: 30-35 months* |  |  |  |  |
| N | 175 | 87 | 88 |  |
| OVERALL | 53.07±0.66 | 53.28±0.6 | 52.87±0.66 | 0.00 |
| Z_OVERALL | -0.64±0.84 | -0.38±0.76 | -0.91±0.84 | 0.00 |
| *Child age: 0-35 months* |  |  |  |  |
| N | 2910 | 987 | 1923 |  |
| OVERALL | 47.26±4.6 | 47.79±4.6 | 46.99±4.58 | 0.00 |
| Z_OVERALL | -0.25±1.13 | -0.18±1.19 | -0.28±1.09 | 0.03 |

